# Epidemiology of COVID-19 vs. Influenza: Differential Failure of COVID-19 Mitigation among Hispanics

**DOI:** 10.1101/2020.09.21.20199018

**Authors:** William E. Trick, Sheila Badri, Kruti Doshi, Huiyuan Zhang, Katayoun Rezai, Michael J. Hoffman, Robert A. Weinstein

## Abstract

**BACKGROUND:** During the early phases of the COVID-19 pandemic, predominantly African-American or Hispanic communities were disproportionately impacted. We sought to better understand the epidemiology of COVID-19 among hospitalized Hispanic patients by comparing individual and census-tract level characteristics of patients diagnosed with COVID-19 to those diagnosed with influenza, another viral infection with respiratory transmission. We evaluated the temporal changes in epidemiology across race-ethnicity related to a shelter-in-place mandate.

**METHODS:** We evaluated patients hospitalized at Cook County Health, the safety-net health system for the Chicago metropolitan area. Among self-identified hospitalized Hispanic patients, we compared those with influenza (2019-2020 influenza season) to COVID-19 infection during March 16, 2020 through May 11, 2020. We used multivariable analysis to identify differences in individual and census-tract level characteristics between the two groups.

**RESULTS:** Relative to non-Hispanic blacks and whites, COVID-19 rapidly increased among Hispanics during promotion of social-distancing policies. Whereas non-Hispanic blacks were more likely to be hospitalized for influenza, Hispanic patients predominated among COVID-19 infections (40% relative increase compared to influenza). In the comparative analysis of influenza and COVID-19, Hispanic patients with COVID-19 were more likely to reside in census tracts with higher proportions of residents with the following characteristics: Hispanic; no high school diploma; non-US citizen; limited English speaking ability; employed in manufacturing or construction; and, overcrowding. By multivariable analysis, Hispanic patients hospitalized with COVID-19 compared to those with influenza were more likely to be male (adjusted OR=1.8; 95% CI 1.1 to 2.9) or obese (aOR=2.5; 95% CI 1.5 to 4.2), and to reside in a census tract with ≥40% of residents without a high-school diploma (aOR=2.5; 95% CI 1.3 to 4.8).

**CONCLUSIONS:** The rapid and disproportionate increase in COVID-19 hospitalizations among Hispanics after the shelter-in-place mandate indicates that public health strategies were inadequate in protecting this population. In particular, those residing in neighborhoods with lower levels of educational attainment.

## INTRODUCTION

There have been many noteworthy contributions to the literature on the epidemiology of COVID-19 including comparisons of the burden of cases and mortality by race-ethnicity. During most of 2020, the U.S. emerged as the country with the highest absolute number of cases and deaths.^1^ It is now well recognized that the dramatic increase in transmission after the first few months of the pandemic in the U.S. occurred disproportionately among non-Hispanic black and Hispanic communities.^2^ Factors highlighted as potential drivers of this early and sustained transmission include overcrowding and limited ability to work remotely leading to challenges in social distancing, multigenerational families, and a high-prevalence of comorbidities.^3-5^

In the Cook County Health system, which provides care for the most vulnerable populations of the Chicago metropolitan region, also predominantly non-Hispanic blacks and Hispanics, we noted an early surge in admissions of Hispanic patients. We sought to better understand the epidemiology of COVID-19 in the Hispanic community by comparing patient- and census tract-level factors of patients hospitalized due to COVID-19 to those of patients hospitalized due to influenza infection, another viral infection with respiratory transmission, which historically has impacted non-Hispanic blacks with rates at least as high, if not higher, than Hispanics.^6-8^ We evaluated temporal trends in COVID-19 by race-ethnicity and evaluated temporal changes in the context of social distancing policies and their resultant impact on mobility.

## METHODS

We assembled a cohort of patients hospitalized for influenza during the 2019-2020 influenza or COVID-19 during the early phases of the pandemic, through May 11, 2020. We identified patients from a research data warehouse, which contains clinical data on all health system patients; ecological variables are captured through routine address cleaning and geocoding with linkage to U.S. census data.^9^ We restricted the cohort to laboratory-confirmed influenza and COVID-19 cases; i.e., inclusion in the COVID-19 cohort only included hospitalized patients with SARS-CoV-2 detected by polymerase chain reaction—our first detected case was March 16, 2020. We focused on inpatients because persons hospitalized for respiratory infection during the influenza season routinely are evaluated for influenza; in contrast, laboratory confirmation of influenza in the emergency department is much less common.

For patient-level characteristics, we used self-identified race-ethnicity; patient co-morbidities were captured through ICD-10 diagnosis codes; obesity was defined as a body mass index ≥30 kg/m^2^. Ecological variables were included based on five-year estimates published by the U.S. Census Bureau as the American Community Survey.^9^ We made a priori selections of ecological variables to explore their association with COVID-19 infection and we calculated a Social Vulnerability Index.^10^

We evaluated the prevalence and associations between patient- and ecological-factors for COVID-19 versus influenza infection using bivariable analyses. We constructed multivariable logistic regression models to explore the association between COVID-19 and influenza. Because of multicollinearity between ecological variables, we entered all ecological variables into the final model to explore the strength of relationships with relatively parsimonious models. To more intuitively express the quantitative association between census tract variables and COVID-19 infection, we constructed a final model with dichotomous categorization of census tract variables into upper quartile vs the lower three quartiles. To evaluate temporal trends across racial and ethnic groups, we segmented calendar years into weeks and calculated the proportion of hospitalization for Hispanics, non-Hispanic Blacks, and non-Hispanic whites. We constructed graphs over time using locally smoothed polynomial regression plots with 95% CI bands. All analyses were performed using Stata software, version 14.2.

## RESULTS

Among Hispanics, relative to hospitalizations for influenza infection during the entire 2019-2020 influenza season (n=115), there were over two-fold more hospitalizations due to COVID-19 infection (n=278) during our ∼two-month study period. Compared to influenza, patient admissions for treatment of COVID-19 infection were much more likely to be Hispanic compared to those admitted for influenza, (59% vs 42% respectively; P<0.001), Figure. The relative increase in COVID-19 infection among Hispanics became apparent by the third week after the initial COVID-19 hospitalization and continued to increase until reaching a plateau of over 50% of all patient admissions during Week 5, Figure.

**Figure.**
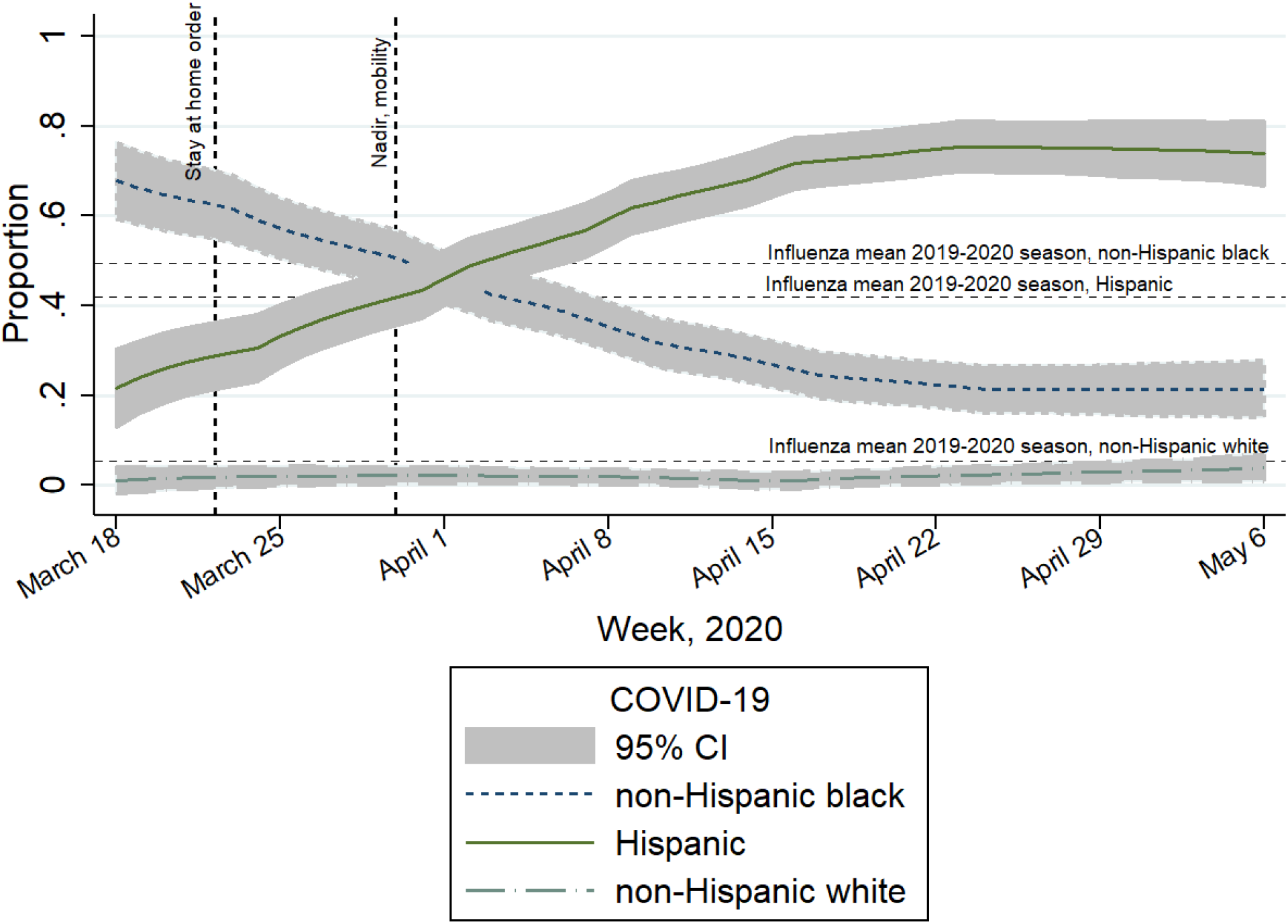
Weekly trend in hospital admissions by race-ethnicity for COVID-19 compared to the mean race-ethnicity values for influenza during the 2019-2020 influenza season.^a^ Findings presented in the context of social control policies and their resultant impact on mobility (represented by vertical lines), Cook County Health, Illinois. ^a^There was a 42% relative increase in the proportion of hospitalized patients who were Hispanic compared to influenza infection.

When we evaluated patient-level factors associated with COVID-19 compared to influenza among hospitalized Hispanic patients, COVID-19 patients were more likely to be male or obese, and less likely to have a diagnosis of asthma, chronic obstructive pulmonary disease (COPD), or heart failure, Table 1. Regarding patient outcomes, COVID-19 infected patients were more likely than those who acquired influenza to require admission to an ICU, and much more likely to require mechanical ventilation or die, Table 1.

**Table 1.**
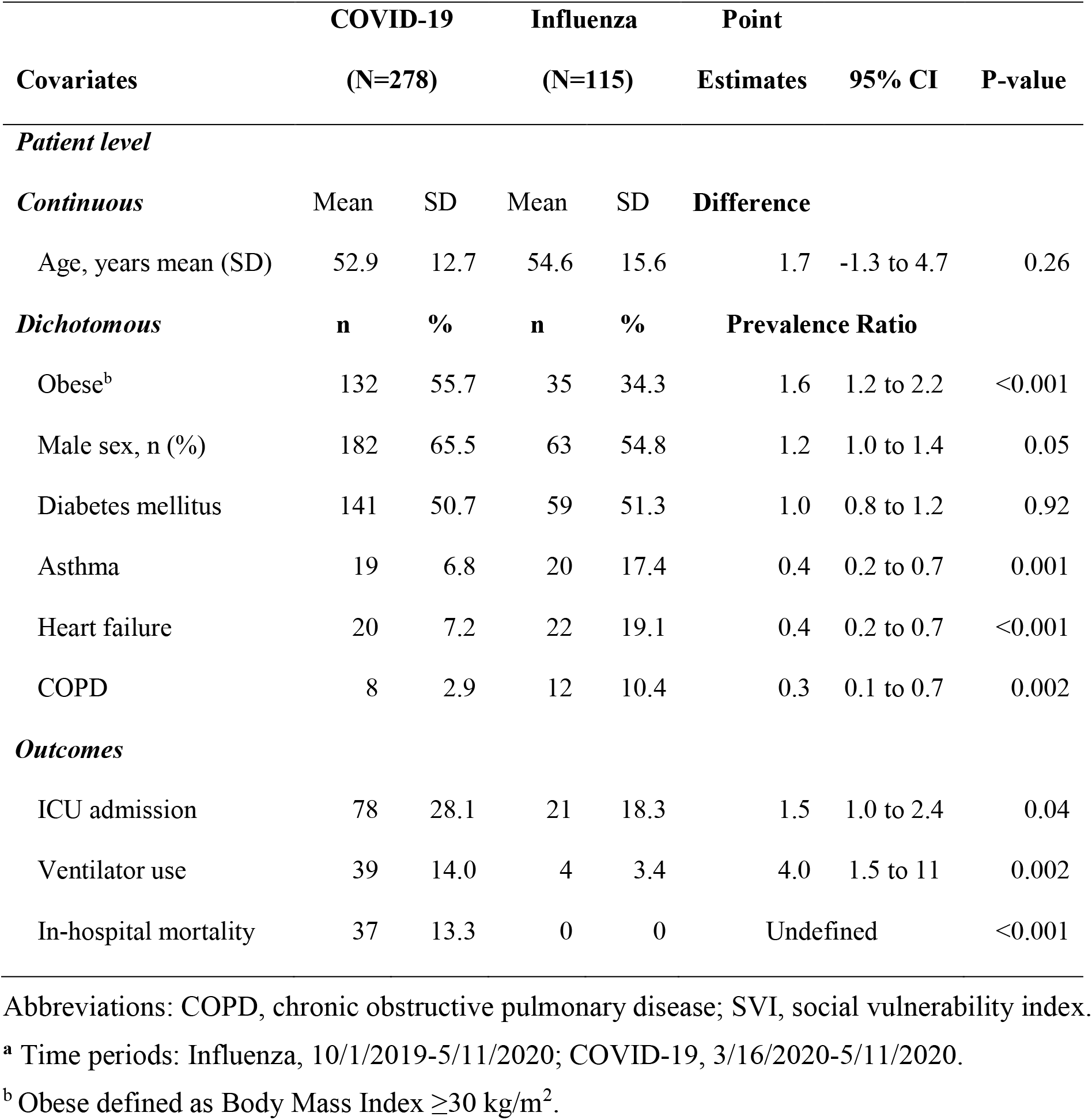
Comparison of patient-level characteristics among hospitalized Hispanic patients with COVID-19 versus influenza infectiona, Cook County Health, Chicago, IL.

Among hospitalized Hispanic patients, ecological factors significantly associated with COVID-19 compared to influenza infection were a higher proportion of the census tract population being the following: Hispanic; Spanish language preference; not U.S. citizens; no high-school diploma; working in manufacturing or construction; and, overcrowding, Table 2.

**Table 2.**
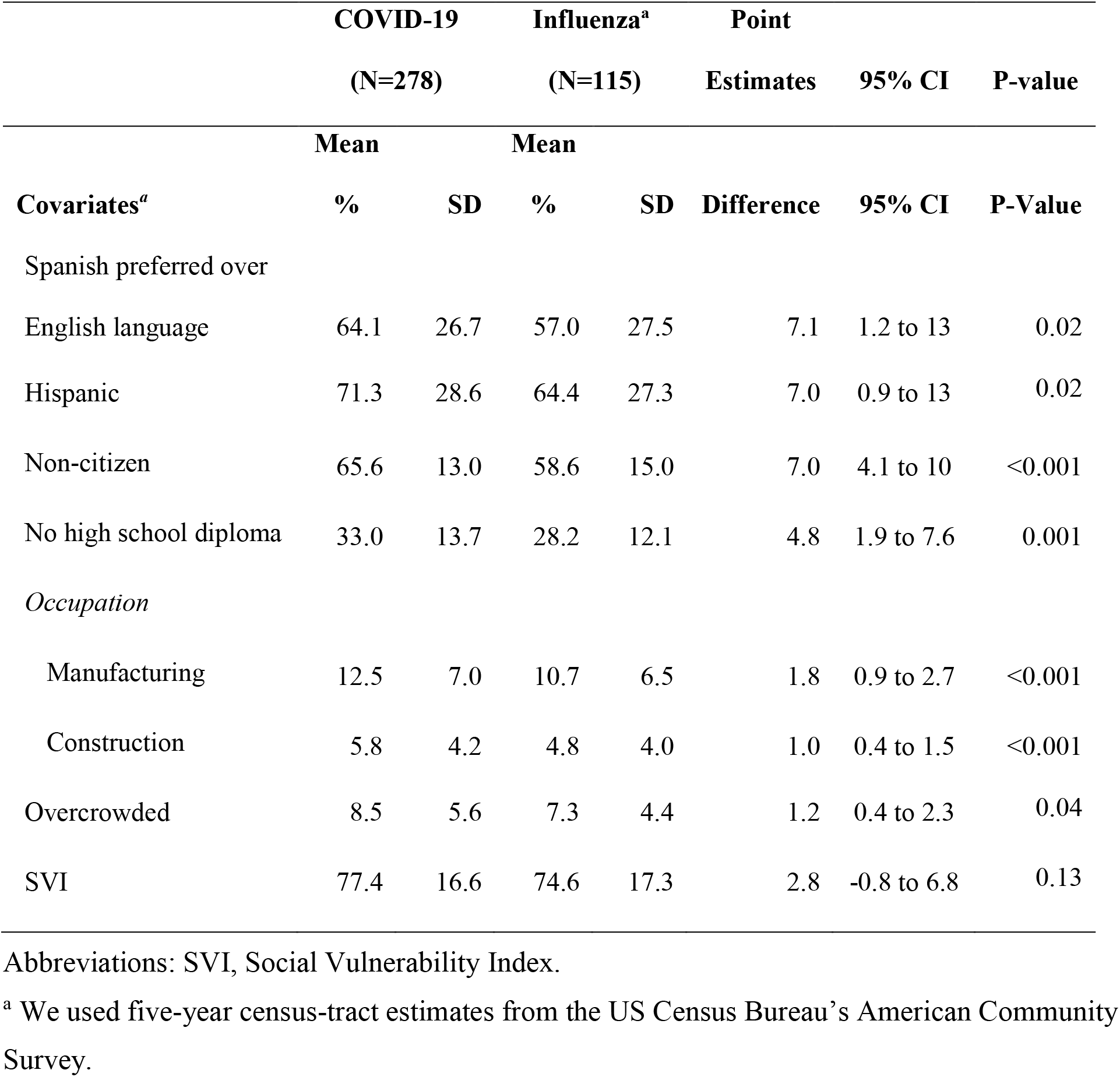
Comparison of neighborhood characteristics of hospitalized Hispanic patients with COVID-19 versus influenza infection, Cook County Health, Chicago, IL.

By multivariable analysis, COVID-19 patients were more likely to be obese, male, or reside in a census tract where ≥40% of residents (upper quartile) reported no high-school diploma; and, less likely to have asthma or heart failure, Table 3. Variable selection during multivariable model development was complicated by multi-collinearity among the ecological variables from census data, e.g., we found moderate to strong correlations between preference for Spanish-language and having no high-school diploma (correlation coefficient [rho]=0.81), being a non-citizen (rho=0.60), and employment in manufacturing (rho=0.73) or construction (rho=0.51).

**Table 3.** Evaluation of ecologicial- and patient-level characteristics by multivariable analysis, COVID-19 versus influenza infectiona. Hispanic patients hospitalized at Cook County Health, Chicago, IL.

| **Variable** | **aOR** | **95% CI** | **P-value** |
| --- | --- | --- | --- |
| Obese | 2.5 | 1.5 to 4.2 | <0.001 |
| Census tract, $\geq 40\%$ without HS diploma<sup>b</sup> | 2.5 | 1.3 to 4.8 | 0.007 |
| Male | 1.8 | 1.1 to 2.9 | 0.03 |
| Asthma | 0.4 | 0.2 to 0.9 | 0.02 |
| Heart failure | 0.3 | 0.2 to 0.7 | 0.001 |

Abbreviations: HS, High School
<sup>a</sup> Time periods: Influenza, 10/1/2019-5/11/2020; COVID-19, 3/16/2020-5/11/2020.
<sup>b</sup> Upper quartile for all census tracts represented by the study population.

## DISCUSSION

By comparing COVID-19 to influenza infection, we confirmed previous reports that Hispanic patients experienced a disproportionate burden of COVID-19.^11^ During the early phases of the pandemic, as social controls were promoted in the Chicago region through the local and state departments of public health, we observed a dramatic and sustained increase in the proportion of COVID-19 infections among Hispanics. The dramatic increase was temporally associated with Illinois’ stay-at-home guidance and the closure of non-essential businesses enacted March 21, 2020. Associated with these policies was a resultant decrease in overall population mobility—a nadir of 56% reduced mobility was attained by March 29, 2020.^12^

The impact of social-distancing interventions would be expected to be effective primarily for individuals with capacity to “shelter-in-place”, while transmission likely would continue unabated—possibly even accelerating—for individuals who were unable to “shelter-in-place” due to on-site employment or reduced job mobility due to legal status, occupation, and reduced opportunities associated with lower levels of educational attainment.^13,14^ Certain occupations have been associated with increased risk of COVID-19 infection and although we didn’t collect individual patient occupation, our ecological analysis identified that among hospitalized Hispanic patients, COVID-19 was more likely than influenza infection to occur among residents of census tracts with a higher reported proportion of employment in construction and manufacturing. Additionally, the sociospatial characteristics, such as intense social ties, and multi-generational households that were described as protective for Hispanic communities during Chicago’s 1995 heat wave, may have played a role in transmission of COVID-19 among social contacts in Hispanic communities outside of work.^15^

Compared to influenza and not surprising, COVID-19 patients were more likely to have severe infection as manifested by a higher likelihood of ventilator use and mortality. The dramatic mortality difference is consistent with estimates of increased mortality risk from COVID-19.^16^ Although mortality could have been influenced by a considerable increase in the stress on the health system, the Chicago region, including Cook County Health, never experienced a period of time in which there were no available ICU beds or ventilators.

The increased risk of infection for males and for obesity has been described for both influenza and COVID-19;^17,18^ however, these two characteristics were more common among COVID-19 compared to influenza patients. Our finding a lower prevalence of asthma or heart failure among hospitalized patients with COVID-19 infection should not be interpreted as an indicator that individuals with these chronic conditions are not at increased risk for COVID-19 compared to the general population. Rather, this finding may be explained by the higher virulence of COVID-19 compared to influenza; i.e., COVID-19 is more likely to result in illness severe enough to require hospitalization independent from pre-existing pulmonary or cardiac conditions.

In summary, in a health system that cares for patients who come from the most socially vulnerable communities, COVID-19 disproportionately affected Hispanic individuals and communities; a tragic situation that was foretold.^19^ This suggests a differential and importantly disparate impact of mitigation measures across communities, even among Hispanic patients. Additional studies are needed to better understand individual-level behaviors among Hispanics during the COVID-19 pandemic. For the current and future pandemics we need anticipatory plans that transcend community and individual risks, without which, disparities will result from well-intentioned public health policies and interventions.

## Data Availability

We have not made the data publicly available. It is an unfunded project with patient privacy concerns that would complicate providing a publicly-accessible dataset.

## ACKNOWLEDGEMENTS

There were no external funds for this evaluation. The authors have no conflicts of interest to report. We acknowledge Vanessa Sarda for her guidance on data analysis and presentation, and drafting of the manuscript.

